# A customizable calculation tool for allocation of adrenal vein sampling in primary aldosteronism in diverse populations

**DOI:** 10.64898/2026.02.05.26345289

**Authors:** AA Leung, SJ Przybojewski, M Klamrowski, C Caughlin, C Wright, JL Pasieka, VC Wu, AYH Lin, CH Tsai, CC Chang, G Hundemer, J King, K Austin, K Mellor, X Hu, J Low, JJ Burkart, GA Kline

**Author notes:** Corresponding author and reprint requests: Gregory A. Kline MD FRCPC, Clinical Professor of Medicine, University of Calgary 1820 Richmond Rd SW, Calgary, AB, Canada T2T 5C7 T, F, E.

## Abstract

**Background:** Primary aldosteronism(PA) screening is recommended but disease prevalence exceeds the availability of adrenal vein sampling(AVS).

**Methods:** An AVS optimal allocation tool for health systems was developed using administrative data and AVS registries from Calgary and Taiwan. Four easily-definable phenotypes of PA based on an elevated aldosterone-renin-ratio (ARR), and the presence/absence of hypokalemia or adrenal mass were identified, representing progressively severe PA and stepwise increasing rates of AVS-defined lateralization.

Using supply-and-demand principles, a customizable, web-based tool was developed that considers PA referral volume, PA phenotype prevalence, maximum AVS available/year, AVS success rate, and desired rate of finding unilateral disease.

**Results:** The most prevalent phenotype of PA was characterized by an elevated ARR and hypokalemia but no adrenal mass (41.9 [39.9–43.9]%); hypokalemia and adrenal mass accounted for (15.6[14.4-16.9]%) of cases. There was a progressive increase in AVS lateralization rate with increasing severity of phenotype observed in both the Calgary and Taiwan data, ranging from (20-39%) in those with PA without hypokalemia or adrenal mass to (70-90%) in those with hypokalemia and adrenal mass. After accounting for institution-specific lateralization rates and allowing for system-level differences in high- and low-volume PA referrals, and high- and low AVS availability, the customizable AVS allocation tool was able to generate individualized strategies ranging from restrictive (exclusive reservation of AVS for cases with hypokalemia and adrenal mass) to more inclusive strategies (assigning a proportion of AVS allocation to less severe PA cases).

**Conclusions:** An AVS allocation tool that uses common, simple, and globally available PA case data may assist in health system AVS program case allocation for maximum equity and wait-list control.

**GRAPHICAL ABSTRACT:** 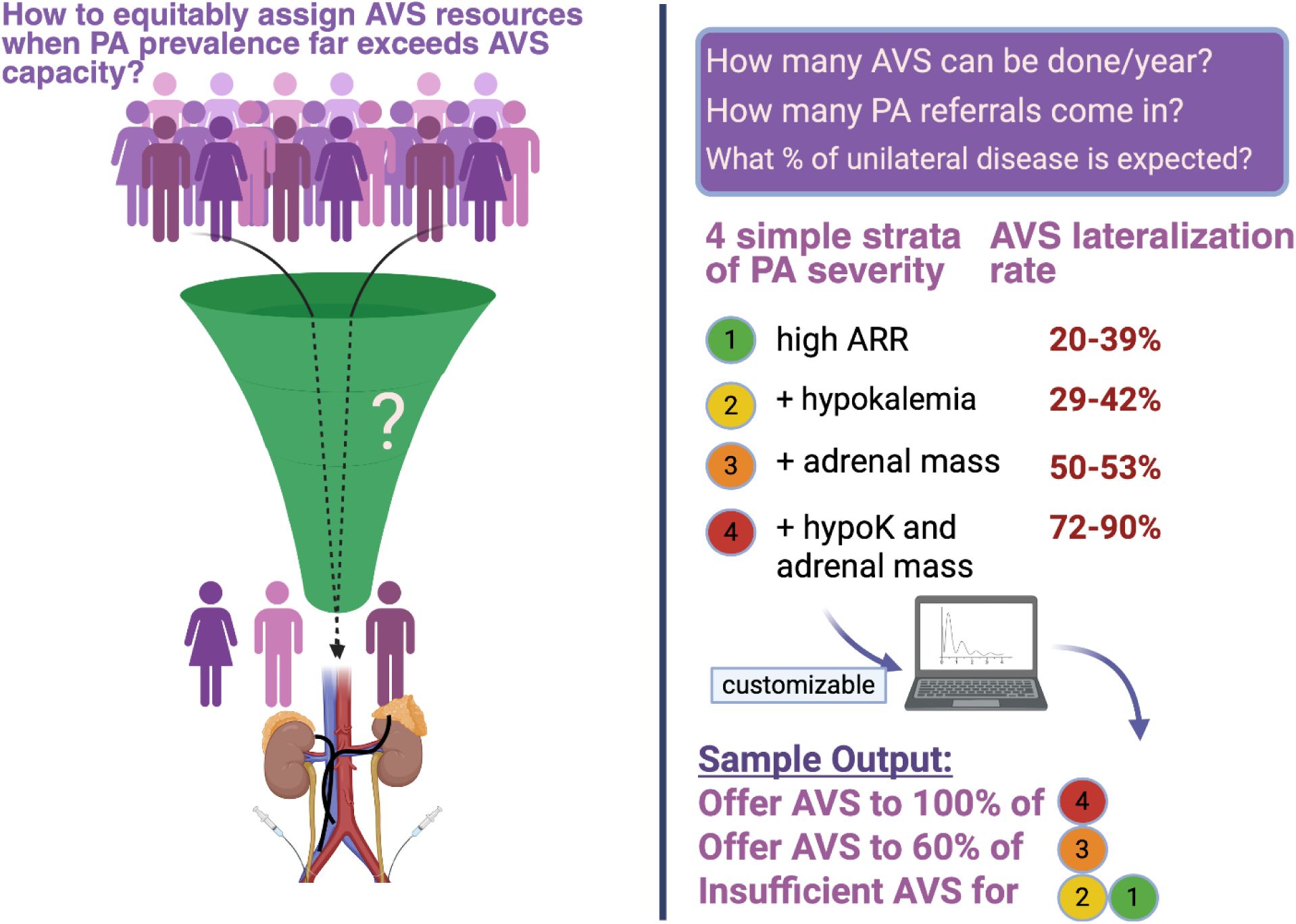

## Introduction

Primary aldosteronism (PA) is a common form of hypertension with an increased risk of morbidity and mortality compared to primary hypertension(1)(2)(3). Previously believed to be rare, it is now recognized as being present in 11-22% of hypertension cases(4), sufficient to warrant universal biochemical screening in hypertensive patients, as recommended by multiple international guidelines(5)(6).PA is operationally subtyped into forms believed to arise from a single (unilateral) adrenal gland and those from multiple (bilateral) adrenal sources. Differentiation is most commonly through abdominal computed tomography (CT) imaging and adrenal vein sampling (AVS). A key distinction in PA is whether the condition lateralizes to one adrenal gland or is bilateral as this guides definitive management. While medical treatment with MRAs are recommended for patients with bilateral PA, surgical adrenalectomy is recommended(5) for patients with lateralized PA and results in high rates of clinical cure, biochemical cure, and most importantly improved longitudinal health outcomes (well beyond what is achieved with medical therapy)(7)(8). Despite numerous calls for wider PA screening(9)(10) screening rates remain low with studies from multiple countries suggesting that no more than 1-2% of affected patients are ever diagnosed and treated(11)(12)(13)(14)(15). However, given the very high prevalence of hypertension(16)(5), it is anticipated that implementation of recent guideline PA screening recommendations will likely result in an enormous number of positive screening tests.

Adoption of universal screening for PA will have far-reaching implications on resource utilization and healthcare delivery with the potential of affecting millions of people globally(17). The greatest bottleneck is expected to be with subtyping. Universal screening for PA may lead to an unsustainable surge in cases, creating challenges in already overburdened healthcare systems with limited access to subtyping investigations, such as AVS(18). The 2025 Endocrine Society guidelines(5) implicitly recognize the potential resource implications of universal screening and have suggested a conditional recommendation to perform aldosterone suppression testing in most patients with PA to limit the number of people who will be referred for AVS. However, this strategy does not accurately differentiate patients who have unilateral (surgical) PA from those with bilateral (medical) disease(19)(20). There are prior studies which aimed to generate multivariable prediction models to predict whether a PA patient has unilateral or bilateral adrenal disease (refs). Regardless of model performance, this approach is only moderately helpful because it only modulates the “demand” side of a supply-and-demand situation. There are unlikely to be any health systems in the world where AVS is completely unlimited and thus the “supply” side of the equation cannot be ignored. As such, the issue cannot be completely solved by predicting AVS results but rather, AVS allocation needs to be addressed.

Therefore, a validated AVS allocation selection strategy is urgently needed to address the anticipated surge in cases and to inform policy. Addressing this, we developed an AVS allocation tool that can be easily customized for any institution to effectively select patients who would benefit most from AVS according to the expected number of patients with PA, their basic clinical characteristics, and the maximum number of AVS procedures that can be accommodated per year.

## Methods

The study was approved by the Conjoint Health Research Ethics Board at the University of Calgary. A waiver of consent was granted for access to personal identifiable health information consistent with the conditions of section 50 of the Health Information Act of Alberta, Canada.

### Patient Population and Data Sources

We previously assembled a retrospective, population-based cohort (CALCTI) of patients with PA in the province of Alberta, Canada(21). This served as our study’s “derivation cohort” where we initially examined our AVS allocation tool. This cohort was assembled by identifying patients through linked administrative health databases of Alberta Health (a government ministry that provides universal healthcare coverage to more than 99% of residents), which included demographic, laboratory, and diagnostic imaging data. We identified 1.1 million residents of Alberta who were 18 years or older between April 1, 2012, and July 31, 2019, who had hypertension using a validated case definition (based on 1 hospitalization or 2 physician claims), as previously described(11)(22). Among those with hypertension, we identified all patients who had an elevated aldosterone-renin-ratio (ARR; consistent with PA) and linked their records with the regional AVS database based in Calgary, Alberta, Canada, which captured over 99% of all AVS procedures performed in the province(23). Using this data, we were able to estimate the prevalence of PA among those with hypertension who were screened and calculate the frequency of lateralizing disease among those who received AVS at our centre.

### Predictors of Lateralization

Apart from having an elevated ARR, the presence of hypokalemia and an adrenal mass have been described as strong clinical predictors for lateralization with AVS, as we and others have previously shown(24)(25)(26). These variables are commonly reported, routinely collected, and easily understood. Using these variables, we described 4 categories of PA for our AVS allocation tool. All patients were confirmed to have hypertension, low renin, and non-suppressed aldosterone, as are sine qua non for PA(27)(28). Patients with PA were categorized into one of the four following groups (with ascending probability of lateralization from CAL1 to CAL4):

CAL1: No hypokalemia, no adrenal mass

CAL2: Any Hypokalemia (<3.5 mmol/L), no adrenal mass

CAL3: Adrenal mass >0.7 cm, no hypokalemia

CAL4: Hypokalemia and adrenal mass > 0.7 cm

### Adrenal Vein Sampling

Lateralization was determined using AVS. The decision to perform AVS was a shared decision between the physician and patient. Until recently, the demand for AVS had not surpassed its availability in Calgary, permitting broad access to AVS when it was deemed to be important for management decisions. On average, over the last decade, we performed approximately 80-100 AVS procedures per year(23). AVS was performed pre and post cosyntropin bolus and infusion as previously described(29); for the purpose of this analysis, all AVS results were retrospectively analyzed in accordance with the Endocrine Society 2025 AVS interpretation guidelines where pre-cosyntropin success was defined as a cortisol selectivity index > 2 and post-cosyntropin selectivity index > 5. Lateralization was defined as a lateralization index > 4 at either pre or post cosyntropin sampling(5).

### AVS Allocation Tool

Using the four categories (CAL1-CAL4) described above, we created an AVS allocation tool that would be adaptable to any setting. The tool was made to be customizable, such that the overall prevalence of PA (and the proportion of patients with PA that would be classified into any of the four predefined CAL1-CAL4 categories), and locally derived estimates of the expected rates of lateralizing PA within each of the four CAL1-CAL4 categories could be easily adjusted. This permitted customization of the tool based upon local referral population characteristics and differing AVS protocols and interpretive criteria.

The AVS allocation tool outputs were developed to optimize AVS referrals (i.e. maximize the number of patients referred for AVS with a high probability of lateralization) in consideration of local resources (i.e. maximum number of AVS available per year) according to the predefined CAL1-CAL4 categories. In addition, we considered that third party/government payors may require a certain proficiency and outcome from each year’s set of AVS procedures (i.e., the minimum acceptable rate of detecting unilateral PA among all AVS performed). For instance, health systems with very limited AVS resources may require a very high overall rate of lateralization, to maximize the number of surgical cases detected (i.e. prioritizing a high specificity for unilateral PA). On the other hand, health systems that can accommodate a large volume of AVS may be willing to accept a lower rate of lateralization (i.e. prioritizing a high sensitivity for unilateral PA).

### Statistical Analysis

We used the derivation cohort to estimate the overall proportion of patients with PA who would be categorized as CAL1-CAL4 and the associated lateralization rates with AVS for each category. Then, based on the number of patients referred for the assessment of PA, expected lateralization rates, and AVS resource availability, we calculated the maximum number of PA cases that could be referred per year for patients in each of the CAL-CAL4 categories. In addition, we calculated the predicted program lateralization rate and provided a decision box to indicate whether the outputs satisfied the minimum acceptable lateralization rate/number of AVS available. Finally, the calculator output was built to include an estimate of the percentage of CAL1-CAL4 groups who could be offered AVS under the desired program parameters.

We included an optional component to address the fact that the AVS allocation calculator unrealistically assumes a perfect rate of AVS catheterization success by default. If an AVS operator had less than 100% technical catheterization success, this would add a number of non-diagnostic cases that nonetheless use up AVS resources. Should such cases undergo repeat AVS, that would require a second AVS spot for the same patient. Therefore, the AVS allocation calculator was designed to also permit an additional input so that the local AVS technical success rate could be adjusted, with subsequent updated total number of AVS potentially required if each failed AVS attempt were to be repeated.

A key part of the calculator was that which pertained to the stepwise increase in rate of lateralization with AVS from CAL1 to CAL4. To demonstrate generalizability of our AVS allocation tool and reproducibility of the stepwise increase in lateralization rates with each ascending CAL category, it was reapplied to an independent cohort (TAIPAI) from Taiwan using data collected between November 2023 and December 2025 and AVS interpreted according to the 2025 Endocrine Society guidelines. The TAIPAI cohort has been previously validated in multiple studies of PA, AVS and treatment outcomes(30)(31)(20). Additionally, to show that our AVS allocation tool can be customized to accommodate a wide variety of AVS interpretation criteria that may be used around the world, we performed a sensitivity analysis of the Calgary cohort, using the local Calgary AVS interpretation criteria instead of the 2025 Endocrine Society guidelines, where pre-cosyntropin SI > 2 and post-cosyntropin SI > 3 are considered success; lateralization is any LI > 3. The final calculator has been made freely available online (https://avs-utilization-calculator.vercel.app/). The calculator was created as a web-interface and statistical analysis with MedCalc 19.0.7 (Ostend, Belgium), p< 0.05 taken as the level of statistical significance.

## Results

Patients with PA were categorized according to the CAL1-CAL4 groups, as summarized in Table 1. The total number of patients screened for PA was 4093, and the estimated population prevalence (95% confidence interval) of normokalemic PA without adrenal mass (CAL1) was 32.3% (30.6-34.1), those with hypokalemic PA but no mass (CAL2) was 41.9% (39.9-43.9), and those with hypokalemic PA and adrenal mass (CAL4) 15.6% (14.4-16.9). PA with adrenal mass but no hypokalemia (CAL3) was less common, seen in 10.1% (9.1-11.1) of the screened and imaged population.

**Table 1:** Population prevalence of CAL 1-4 groups with corresponding AVS lateralization rates (Calgary and TAIPAI)

| CAL group | Population prevalence estimate (CALCTI) in Calgary, 95% CI | Observed AVS lateralization rate by pre- **or** post-cosyntropin sampling in Calgary, using ES2025 guidelines interpretation, total AVS=828 | Observed AVS lateralization rate in TAIPAI, using ES 2025 guidelines interpretation, TAIPAI, total AVS = 151 | Observed AVS lateralization rate by pre- **or** post-cosyntropin sampling in Calgary, using traditional Calgary AVS interpretation* |
| --- | --- | --- | --- | --- |
| CAL 1 | 32.3% (30.6-34.1) | 78/395, 20% | 9/23, 39% | 111/425, 26% |
| CAL 2 | 41.9% (39.9 – 43.9) | 48/163, 29% | 11/24, 46% | 63/163, 39% |
| CAL 3 | 10.1% (9.1 – 11.1) | 195/380, 51% | 28/54, 52% | 238/414, 57% |
| CAL 4 | 15.6% (14.4-16.9) | 165/237, 70% | 45/50, 90% | 179/240, 74% |

\* total evaluable AVS by Calgary selectivity index interpretation scheme = 1242.

From the corresponding CAL1-CAL4 groups within the Calgary AVS database, there was an increasing frequency of lateralization with greater severity of PA as summarized in Table 1. There were 828 unique patients with complete datasets for the present analysis; each patient had pre- and post-cosyntropin data. After excluding pre- or post-cosyntropin results with SI < 2 or < 5 respectively, there were 1175 successful bilateral adrenal samplings for analysis (57% of pre-cosyntropin samples and 92% of post-cosyntropin samples). As expected by association of hypokalemia and particularly adrenal mass with unilateral disease, there was a stepwise increase in the rate of unilateral PA at AVS with increasing CAL1-4 severity. In the CAL 1 and CAL2 groups, unilateral disease was found in 20-29% but rose to 51% in CAL3 and 70% in CAL4 groups. Similar patterns were observed in the TAIPAI cohort. All patients underwent both pre- and post-cosyntropin testing, with unilateral disease detected in 39% and 46% of CAL1 and CAL2 patients, respectively. Lateralization rates further increased to 52% in CAL3 and 90% in CAL4.

Table 2 shows simulations of local applications within a theoretical high AVS resource system and theoretical low-resource system while accounting for the possibilities of both high and low numbers of PA referrals.

**Table 2:** Simulations: High vs. low AVS resource settings with many or few PA referrals*.

| Local parameter | High AVS resources |  | Low AVS resources |  |
| --- | --- | --- | --- | --- |
|  | High number of PA referrals | Low-moderate number of PA referrals | High number of PA referrals | Low-moderate number of PA referrals |
| Number of PA referrals/year* | 1500 | 500 | 1500 | 500 |
| Maximum number of AVS available/year | 360 | 360 | 50 | 50 |
| Minimum acceptable rate of unilateral PA found by AVS | 25% | 25% | 40% | 40% |
| CAL 1 group proportion** | 0.7 | 0.5 | 0.7 | 0.5 |
| CAL 2 group proportion | 0.15 | 0.3 | 0.15 | 0.3 |
| CAL 3 group proportion | 0.05 | 0.05 | 0.05 | 0.05 |
| CAL 4 group proportion | 0.1 | 0.15 | 0.1 | 0.15 |
| Expected number of CAL1 cases | 1050 | 250 | 1050 | 250 |
| Expected number of CAL2 cases | 225 | 150 | 225 | 150 |
| Expected number of CAL3 cases | 75 | 25 | 75 | 25 |
| Expected number of CAL4 | 150 | 75 | 150 | 75 |
| cases |  |  |  |  |
| Number of AVS needed for each allocation strategy and associated program lateralization rate (%) |  |  |  |  |
| CAL4 only | 150 (70%) | 75 (70%) | 150 (70%) | 75 (70%) |
| CAL4 and CAL3 | 225 (63.7%) | 100 (65.3%) | 225 (63.7%) | 100 (65.3%) |
| CAL 4 and CAL3 and CAL 2 | 450 (46.3%) | 250 (43.5%) | 450 (46.3%) | 250 (43.5%) |
| CAL 1-4 (all) | 1500 (27.9%) | 500 (31.8%) | 1500 (27.9%) | 500 (31.8%) |
| Strategy that meets all specified criteria and demand | CAL 3-4 | CAL 2-4 | CAL 4 only | CAL 4 only |
| Total number of AVS potentially needed for chosen strategy and with technical success rate of 91% | 247 | 275 | 164 | 82 |
| Number of AVS available to next lowest CAL group after chosen strategy filled*** | 113 (50% of CAL2 cases) | 85 (34% of CAL1 cases) | none | none |

\*referrals per year within a health system, all providers
\*\*referral proportions customizable; simulation assumes a higher proportion of more hypokalemic/mass cases in an environment where screening is low/targeted to resistant hypertension.
\*\*\*remaining AVS spots that could be used, provided a potentially lower program lateralization rate is acceptable.

Table 3 shows a second simulation using the actual AVS lateralization data from the TAIPAI group. The simulation model demonstrates sensitivity to local PA population features and diagnostic practices.

**Table 3:**
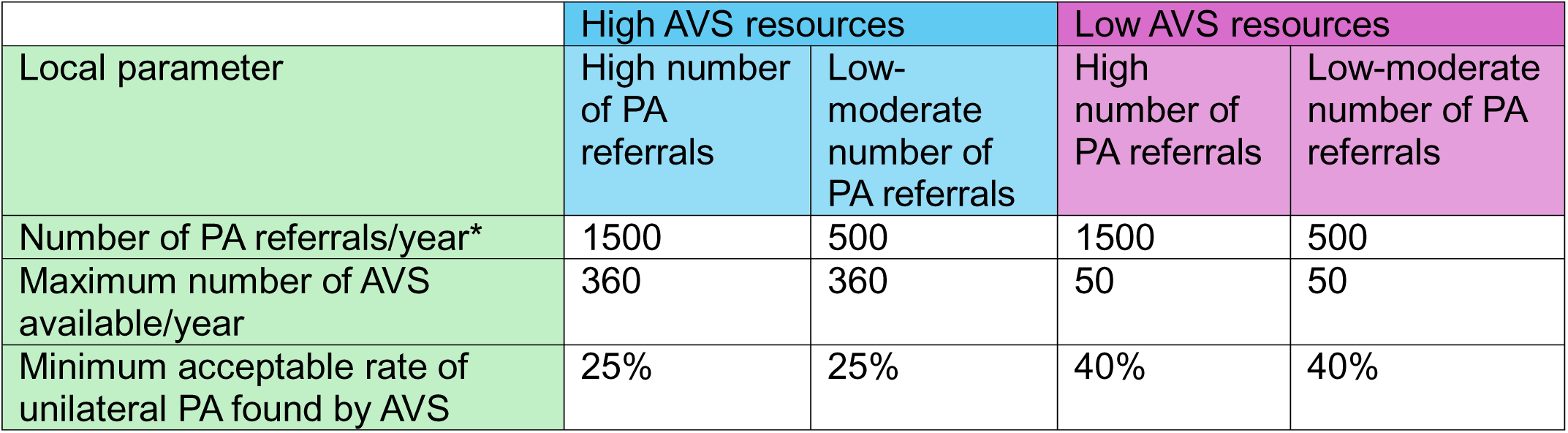
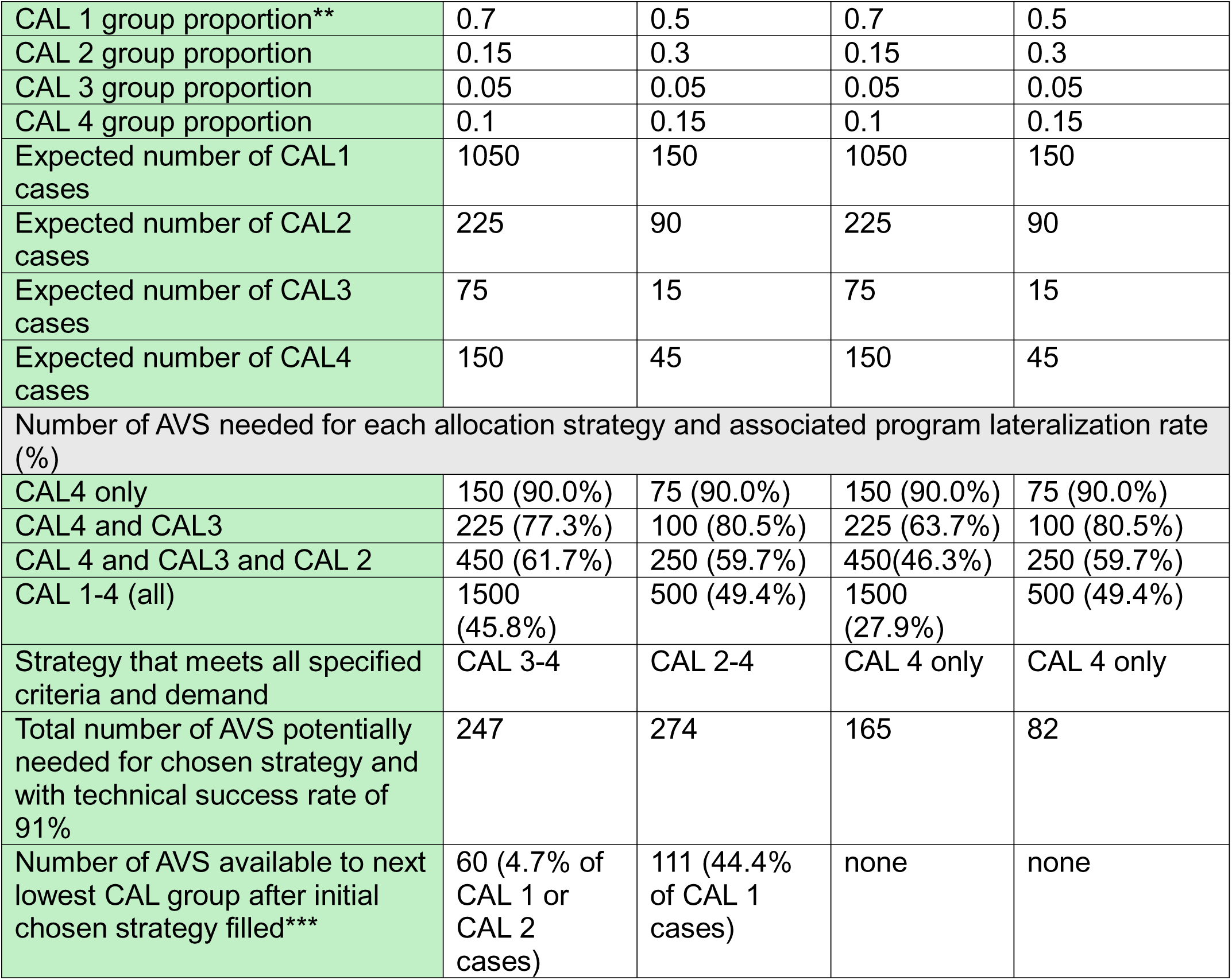
High and Low AVS resources with Many or Fewer PA referrals (TAIPAI model parameters)

## Discussion

We developed an open-access AVS allocation tool that takes into consideration the prevalence of PA and clinically relevant categories that correlate with AVS lateralization rates using simple parameters that are widely available in all practice settings. The calculator is fully customizable and can be used in any health system where differences in PA prevalence, referral patterns, patient volumes, AVS protocols and interpretation schemes, and local AVS availability can be accounted for and accommodated. This calculator is intended to be primarily used at the health system level, rather than at the individual patient level (where the decision of whether to proceed with AVS should be based on shared and informed decision making with patients after considering potential complex clinical context for the latter)(32)(33). However, with integration of supply and demand principles, this calculator may potentially assist decision makers in the development of AVS programs to ensure priority access for those who are most likely to benefit from AVS and to ensure value for investment by setting minimum standards for management-changing findings in a desired percentage of procedures performed. The optimal local data for complete customization of the AVS allocation tool is thus summarized in Supplemental Table 1.

Our use of large sized administrative data and AVS registry results in two large PA specialty centers permits a system-level view of AVS allocation considerations. The Calgary and TAIPAI data need not be assumed to be comparable or generalizable to all settings nor is a specific output validation needed, as the calculator is based upon program design principles and is not intended to necessarily replace PA practice guidelines. However, in view of the disappointing performance of aldosterone suppression tests to select patients for AVS(19)(34)(35), the simplified model presented here may be an attractive option for health systems where unlimited AVS access is not possible. Importantly, our allocation tool does not require specialized additional tests such as metabolomics, hybrid steroid measures, microRNAs (36), or dynamic suppression tests(34), most of which are not available in most parts of the world(18).

Rather the CAL 1-4 groups are based upon data which is easily available (or potentially available) in every case of PA – the presence or absence of hypokalemia and CT adrenal nodule. Both the Calgary and TAIPAI cohorts showed similar step-wise increase in lateralization rates with each incremental category of the CAL 1-4 strata, suggesting these simple characteristics easily identify progressively high-yield AVS categories.

It is important to note that the AVS allocation tool is not based upon nor intended to be validated by surgical outcomes although that could be conducted for quality assurance by any center using it. The AVS allocation tool is intended only to take AVS lateralization rates into account. Subsequent decisions for surgery may be far more nuanced(32)(37)(38) and surgical outcomes determined by a multiplicity of factors which are not relevant to whether AVS was performed or not(39)(40)(41)(42). Surgical outcome prediction rules have been published(43) and may represent an alternative approach to AVS allocation. However, once again, aldosterone suppression tests fail to predict surgical outcomes as well(19)(20) and so other, potentially more complex factors must be considered. A danger of surgical outcome prediction models is the dependence upon short term outcome data which may differ significantly from long term outcome data(44)(45) and the under-valuation of the so-called partial clinical or biochemical response(46)(47) which may fall short of complete cure but still have high relative value to the patient(48)(49).

At the individual patient level, several prediction models have been proposed for both unilateral and bilateral disease. However, most of these models are population-specific, based on very small derivation cohorts, lack quality validation data, or require the collection of more complex biochemical measures that are not easily available in all settings(50)(51)(52)(53). In addition, the existing prediction models are fixed in nature, that is, the user must assume similarity of their patient population to that in the derivation/validation cohort. In reality, different populations and difference PA diagnostic definitions may result in widely variable numbers/proportions of PA cases and PA subtypes(54), emphasizing the importance of having a tool that may be customizable to different populations. Prediction models for lateralization have been shown to be largely non-generalizable(55)(51). Our calculator, based upon principles of supply and demand is not beholden to such limitations, provided reliable local data exist for customization (i.e. for PA prevalence, lateralization rates, etc.). At the least, our calculator provides a framework for those who wish to collect simple, retrospective program audit data for local application. An audit of PA referrals using the CAL1-CAL4 groups would be uncomplicated, as would assessment of matching lateralization rates within an AVS registry. Similarly, our calculator could be used to justify AVS program value to government health payors who may put a high value on procedures that frequently, materially change patient management towards the best outcomes.

A final important feature of the calculator is the ability to determine how many AVS might be available within an institution’s program to serve individual cases that may not fit the minimum standard criteria suggested by the model. For instance, in the high referral/high AVS resource example above (Table 2), even after offering AVS to all patients who are in CAL 4 and CAL 3 categories, there are still 11% of all AVS spots (or 135 in total) that might be offered to patients who do not initially qualify. If AVS access requires gatekeeping, it would be highly useful to both the clinician and the program manager to know how much leeway is available for the inevitable unique individual cases that arise. Knowledge of the number of potential AVS spots not otherwise reserved for the prioritized higher CAL groupings can encourage clinicians to choose wisely, perhaps using some of the other prediction tools suggested.

In similar fashion, an AVS program administrator could use the calculator outputs to determine how many AVS would be required to offer the procedure to each CAL1-CAL4 group. For example, in a system that sees 500 PA referrals per year and where a program lateralization rate of 40% is desired, the health system needs at least 75 AVS spots (6-7 per month) to accommodate just the CAL4 group of patients. To additionally accommodate CAL3, the system would need to offer another 25 AVS spots (8-9 per month for a total of 100 per year). Furthermore, to offer AVS to hypokalemic PA patients without any adrenal mass (CAL 2), the system would need to potentially significantly expand the number of AVS spots available to 250 per year (or 20-21 per month). If scalable, this would still result in an overall lateralization rate of 43.5%, within the value-target set by the system. This too is customizable; some centres may be prepared to offer surgery to CAL 4 patients without AVS as also suggested by recent guidelines(5). By subtracting CAL 4-allocated AVS from the total, these may be re-distributed to lower CAL groups as desired. As functional adrenal molecular imaging for PA subtyping(56) becomes more available, similar allocation considerations may apply; the present tool may be easily adapted to non-AVS modalities in molecular imaging as well.

Key limitations of our tool is that it relies on accurate inputs. Differences in expected vs. actual lateralization rates, and the possibility that referral demographics or AVS protocols change over time may alter the tool’s performance. Moreover, there is inherent subjectivity in selecting the minimum acceptable rate for cases of lateralization. This emphasizes that use of the CAL1-CAL4 AVS allocation tool should not be used as a one-time exercise but rather in the context of ongoing continuous quality improvement of any AVS program. As PA increases in detected prevalence across many countries, this customizable AVS resource allocation tool may prove useful in decision-making for optimal and equitable AVS usage.

## DATA AVAILABILITY

Some or all datasets generated during and/or analyzed during the current study are not publicly available but are available from the corresponding author upon reasonable request and ethical approval.

## Abbreviations

ARR: aldosterone-renin-ratio
AVS: adrenal vein sampling
CT: computed tomography
PA: primary aldosteronism

## Novelty and Relevance

### What is new?

PA guidelines have identified the need for a tool to select PA patients to undergo adrenal vein sampling
We developed a simple PA categorization system using easily accessible common variables that correlate with likelihood of having unilateral disease
We developed an online, customizable tool using the categories in a supply-and-demand model to aid health system prioritization for AVS allocation

### What is relevant?

AVS procedures are not unlimited in availability
Unplanned or inequitable allocation of AVS bookings may cause untenable wait times or de-prioritize patients where AVS is high value
The tool created in this study is customizable to all health systems based on variable supply and demand for PA cases and AVS availability
What are the clinical implications?
This AVS allocation tool permits locally customizable, equitable advance planning for health systems with limited AVS resources
This tool does not require access to experimental, expensive, or restricted tests used in previous AVS prediction rules

**Supplemental Table 1:** Local data recommended for complete customization of AVS allocation tool to population or health system specific needs.

| Data | Suggested source(s) | Sample data from Calgary cohort |
| --- | --- | --- |
| Number of new PA referrals that could potentially be considered for AVS, per year | 1 – Local laboratory data on ARR tests and % above PA screening threshold/year<br>2 – population ICD-10 physician billing codes for primary aldosteronism/year<br>3 – Tertiary hypertension clinic referrals/yr x 30% | Regional, tertiary endocrine hypertension clinic serving population of approx. 1.5M, 600 referrals per year, 80% have PA. |
| Maximum number of AVS procedures potentially feasible within interventional radiology service | Interventional radiology administration | 12-15/month, 180/year |
| AVS catheterization success rate | Local Quality Assurance data | 91% |
| Population prevalence of CAL 1, 2, 3, 4 groups within entire PA referral population | Retrospective review of PA referrals | See main Table 1 |

